# SARS-CoV-2-specific antibody detection for sero-epidemiology: a multiplex analysis approach accounting for accurate seroprevalence

**DOI:** 10.1101/2020.06.18.20133660

**Authors:** Gerco den Hartog, Rutger M. Schepp, Marjan Kuijer, Corine GeurtsvanKessel, Josine van Beek, Nynke Rots, Marion P.G. Koopmans, Fiona R.M. van der Klis, Robert S. van Binnendijk

**Author notes:** **Corresponding author** Gerco den Hartog. **Footnote page**. The authors declare no conflict of interest. The work was supported by the National Institute for Public health and the Environment, The Netherlands.

## Abstract

**Background:** The COVID-19 pandemic demands detailed understanding of the kinetics of antibody production induced by infection with SARS-CoV-2. We aimed to develop a high throughput multiplex assay to detect antibodies to SARS-CoV-2 to assess immunity to the virus in the general population.

**Methods:** Spike protein subunits S1 and RBD, and Nucleoprotein were coupled to distinct microspheres. Sera collected before the emergence of SARS-CoV-2 (N=224), and of non-SARS-CoV-2 influenza-like illness (N=184), and laboratory-confirmed cases of SARS-CoV-2 infection (N=115) with various severity of COVID-19 were tested for SARS-CoV-2-specific concentrations of IgG.

**Results:** Our assay discriminated SARS-CoV-2-induced antibodies and those induced by other viruses. The assay obtained a specificity between 95.1 and 99.0% with a sensitivity ranging from 83.6-95.7%. By merging the test results for all 3 antigens a specificity of 100% was achieved with a sensitivity of at least 90%. Hospitalized COVID-19 patients developed higher IgG concentrations and the rate of IgG production increased faster compared to non-hospitalized cases.

**Conclusions:** The bead-based serological assay for quantitation of SARS-CoV-2-specific antibodies proved to be robust and can be conducted in many laboratories. Finally, we demonstrated that testing of antibodies against different antigens increases sensitivity and specificity compared to single antigen-specific IgG determination.

## BACKGROUND

COVID-19 caused by the newly emerged SARS-CoV-2 resulted in a pandemic in a largely immune-naïve population. The presence of specific antibodies is currently being investigated to assess the induction of an immune response in patients, whilst also to assess the degree of exposure and immunity in the general population [1-3]. As it is a recently emerged coronavirus variant, the kinetics and degree of immunity induced following contact with the virus and COVID-19 disease are largely unknown.

SARS-CoV-2 expresses a Spike protein, highly similar to Spike of SARS-CoV that binds to the Angiotensin converting enzyme 2 (ACE2) [4, 5]. Binding of antibodies to the receptor Binding Domain (RBD) of Spike neutralizes the ability of the virus to infect cells [6]. In addition to elicitation of antibodies to the Spike protein, antibodies are detected against other viral proteins, including Nucleoprotein, N [7]. N is shielded within the virion and therefore N-specific antibodies are probably unable to neutralize the virus. Although N may not be involved in neutralization of the virus, antibodies to N could provide an indicator of exposure to the virus. Antibodies to N induced by SARS-CoV reportedly recognize N of SARS-CoV-2 but not of seasonal coronaviruses [8].

Estimates of the prevalence of seroconversion as proxy for protection of the general population may support health decision making, including the decision to lift lock down measures. To appropriately apply an assay for serosurveys we need to know the precision of the assay, e.g., what is the sensitivity and specificity, which is variable between currently available tests [9, 10]. Performing and sustaining large population studies to assess changing population immunity, demands high-throughput screening assays that are robust and accurate [11]. Many countries now aim to assess the protective status of the general population for COVID-19 using antibody assays. To guarantee high specificity, the assay should be validated with a representative number of sera from patients infected with other coronaviruses and other pathogens causing influenza like illness, but this is often lacking [11-13]. To date, COVID-19 prevalence of seroconverted individuals is relatively low, and there is a risk of significant overestimation if an assay has insufficient specificity (**Table S1**). High specificity is important at this stage (11,12).

Our laboratory has extensive experience in developing multiplex assays to quantify antibodies to many bacterial and viral pathogens in the general population, of which most are part of the national immunization program [1, 14-17]. We developed a high throughput and highly quantitative and specific bead-based multiplex immunoassay to assess the prevalence of sero-positivity in the general population, anticipating the introduction of future SARS-CoV-2 vaccines as well. By multiplexing a broader range of SARS-CoV-2 antigens in one single assay we may generate a better understanding of the proportion of persons that seroconverted. Moreover, in a multiplex assay positivity can be compared between antigens in order to provide a more detailed evaluation of the antibody levels and to enhance assay performance [17]. The assay developed is applied on samples of COVID-19 patients with different severity of disease collected at multiple timepoints to determine the kinetics of seroconversion.

## METHODS

### Serum samples

Serum samples were obtained from the following cohorts: 1) a random selection of individuals (N=224) from a national (Dutch) cohort representing all age groups and obtained 3 years prior to SARS-CoV-2 emergence (Pienter3 study, Netherlands trial register #NL5467); 2) individuals (Table S2) with proven non-SARS-CoV-2 influenza-like illness (hCoV ILI) by coronaviruses (N=110) or other viruses (N=74, Non-hCoV ILI) obtained from RIVM, Bilthoven, The Netherlands, trial register #NL4666 [18], and from Erasmus MC, Rotterdam, collected prior to the SARS-CoV-2 outbreak and at least 2 weeks after PCR detection of the virus; 3) sera of 115 laboratory-(PCR) confirmed COVID-19 patients that were either hospitalized (N=50) or outpatient (N=65) (ErasmusMC and RIVM, medical ethical committee number METC 06/282). Of 73 COVID-19 patients paired samples collected between day 3 and 40 after disease onset were available. Ethical approval was obtained from the Erasmus MC Medical Ethical Committee (MEC-2015-306) to anonymously analyze the used ILI and COVID-19 samples. Informed consent and voluntary informed consent was provided where applicable.

### Assay procedure

The steps in assay validation are similar to recently developed bead-based multiplex immunoassays for CMV, EBV and RSV, with minor modifications as described below [16, 17]. For the multiplex bead-based immune assay the following antigens obtained from Sino Biological were used: SARS-CoV-2 monomeric Spike S1 (40591-V08H) and RBD (40592-V08B), and nucleocapsid (N) (40588-V08B). Microplex fluorescent beads were activated in 50 mM MES pH 5.5. The proteins were diluted to a concentration of 0.2 mg/mL in PBS pH 7.4 and added at 5 µg per 75 µl of activated beads.

An internal reference sample was created by pooling 13 sera of COVID-19 patients with varying IgG concentrations. An arbitrary antibody concentration-unit of ‘100’ was assigned on the basis of the mean fluorescence intensity (MFI) signal in the upper limit of linearity of a 3-fold serial dilution of the reference sample.

25 µl of 1:400 and 1:8000 diluted sera in SM01 buffer (Surmodics) +2% FCS were incubated with antigen-coated beads for 45 minutes at room temperature at 750 rpm in the dark. Following incubation, samples were washed three times with PBS, incubated with PE-conjugated goat anti-human IgG for 30 minutes and washed. Samples were acquired on LX200 or FM3D (Luminex). MFI was converted to arbitrary units (AU/mL) by interpolation from a five-parameter logistic standard curve, using Bioplex Manager 6.2 (Bio-Rad Laboratories) software and exported to MS Excel.

### Assay validation

Different batches of antigen-conjugated beads were incubated with serially diluted sera to test linearity and parallelism between bead conjugations, reference and serum samples. Assay robustness was tested by analyzing a serum panel by 3 different operators on independent days using 2 different bead and/or 2 reference batches. The ability to discriminate IgG concentrations between COVID-19 patients and controls was evaluated by ROC analyses. To select the optimal assay defaults, both the Youden’s J statistic, which allows the selection of settings balancing sensitivity and specificity, and a specificity-optimized cutoff (specificity of at least 98.5% for low prevalence settings of 5-10%) were calculated.

### Data analyses

Data were entered into GraphPad Prism 8.4.1 to generate graphs and perform statistical analyses. For the receiver operator characteristic (ROC) analyses antibody concentrations of cross-sectional Pienter3 participants (N=224), ILI patients with coronavirus (N=74) or other viral infection (N=110) were used as the negative control group and PCR-confirmed COVID-19 samples (N=115) with varying clinical severity were used in the positive group. We selected for serum samples that were obtained more than 10 days post onset of disease symptoms to meet a reasonable degree of seroconversion as shown in recent reports [8, 19]. Both the Youden’s J statistic-determined cutoff and the specificity-optimized cutoff (specificity of at least 98.5%) were determined.

To compare differences in concentrations, data were log-transformed and tested with either a t-test between 2 groups, or One-way ANOVA and Tukey’s multiple comparison test to compare multiple groups and adjusted P-values reported. Antibody kinetics was fitted using a non-linear 4-parameter least square fit in Graphpad Prism 8.4.1.

## RESULTS

### Performance of the assay

We prepared a reference serum by pooling 13 PCR-confirmed COVID-19 sera and tested serial dilutions in the multiplex assay consisting of distinct fluorescent beads coupled to SARS-CoV-2 Nucleocapsid (N), S1 and the S1 subunit RBD (**Figure 1A**). This was repeated for varying batches of beads to assess consistency of performance. The assay was able to quantify concentrations in a 1,000 to 10,000-fold concentration range, using a single dilution of the serum. In order to reliably quantify antibody concentrations between the reference serum and test samples, we confirmed that the reference and a selection of samples display the same rate of decline of fluorescence signal with increasing dilutions, which is referred to as parallelism (**Figure 1B**). These data show that the triplex assay is a highly quantitative assay to detect antibodies to SARS-CoV-2.

**Figure 1.**
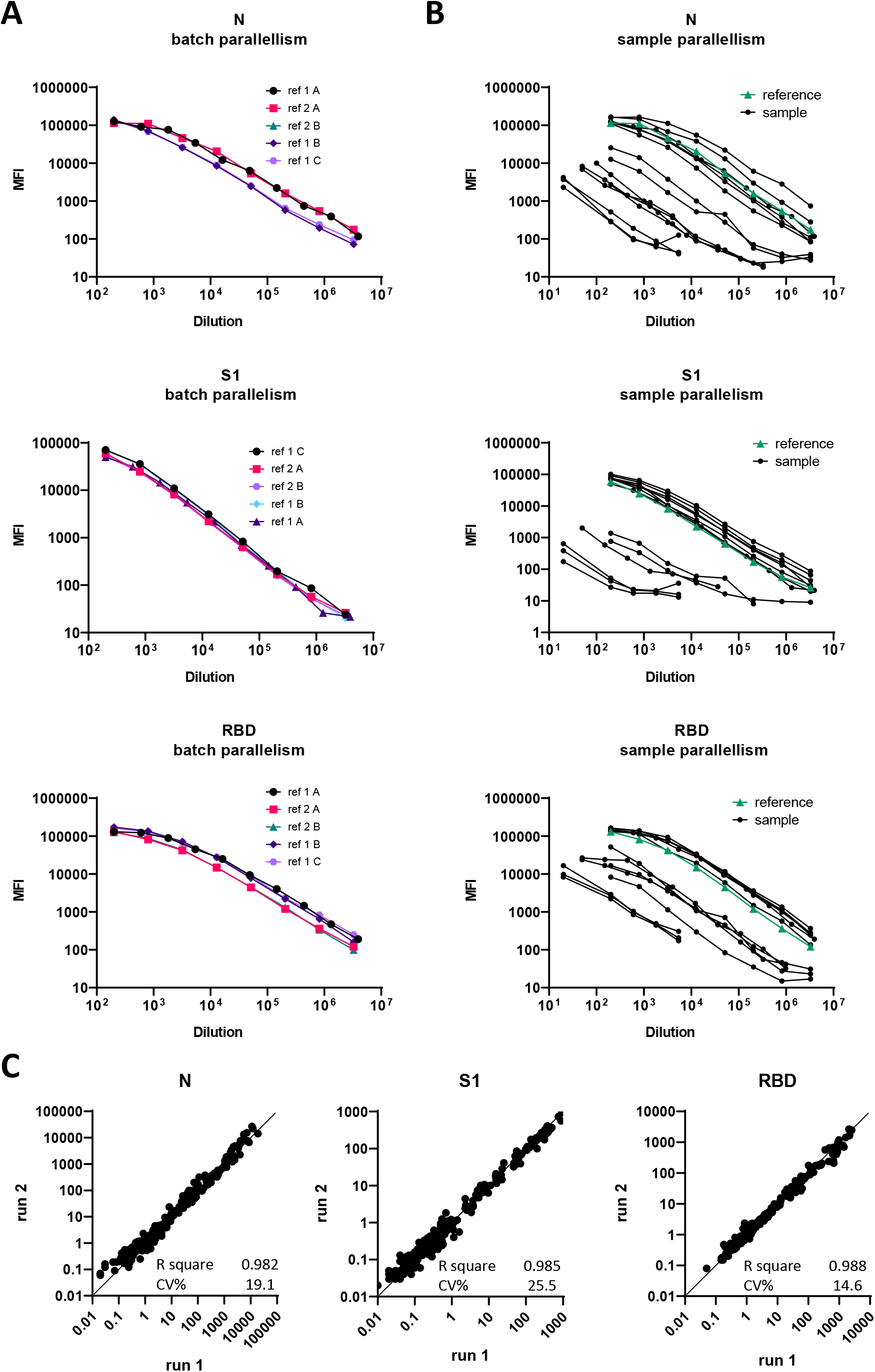
Assay development and validation. **A**. Different lots of antigen-bead conjugations were used to determine the consistency of the mean fluorescence signal of a titrated pooled reference serum. **B**. The reference and serum samples were serially diluted to test parallelism for reliable quantification of antibody concentrations. **C**. Samples were tested in 2 independent runs by different technicians using different bead and reference batches to test robustness of the triplex assay. Concentrations in AU/mL are shown.

Applying an assay in large population and/or longitudinal studies, requires reproducibility of assay results. Therefore, antibody concentrations were determined on two independent days, using a selection of 214 samples for RBD and 268 samples for N and S1 with different concentrations of SARS-CoV-2 antibodies (**Figure 1C**). In addition, the reproducibility test was performed by 3 different technicians using different bead batches and references to reflect the expected maximum variability of the assay over time. Comparison of sample data determined on two independent assays runs resulted in an R square of 0.982, 0.985 and 0.988 for N, S1 and RBD respectively (**Table 1**). The obtained %CVs were 19.1, 25.5 and 14.6 for N, S1 and RBD respectively showing that assay results are reproducible.

**Table 1.**
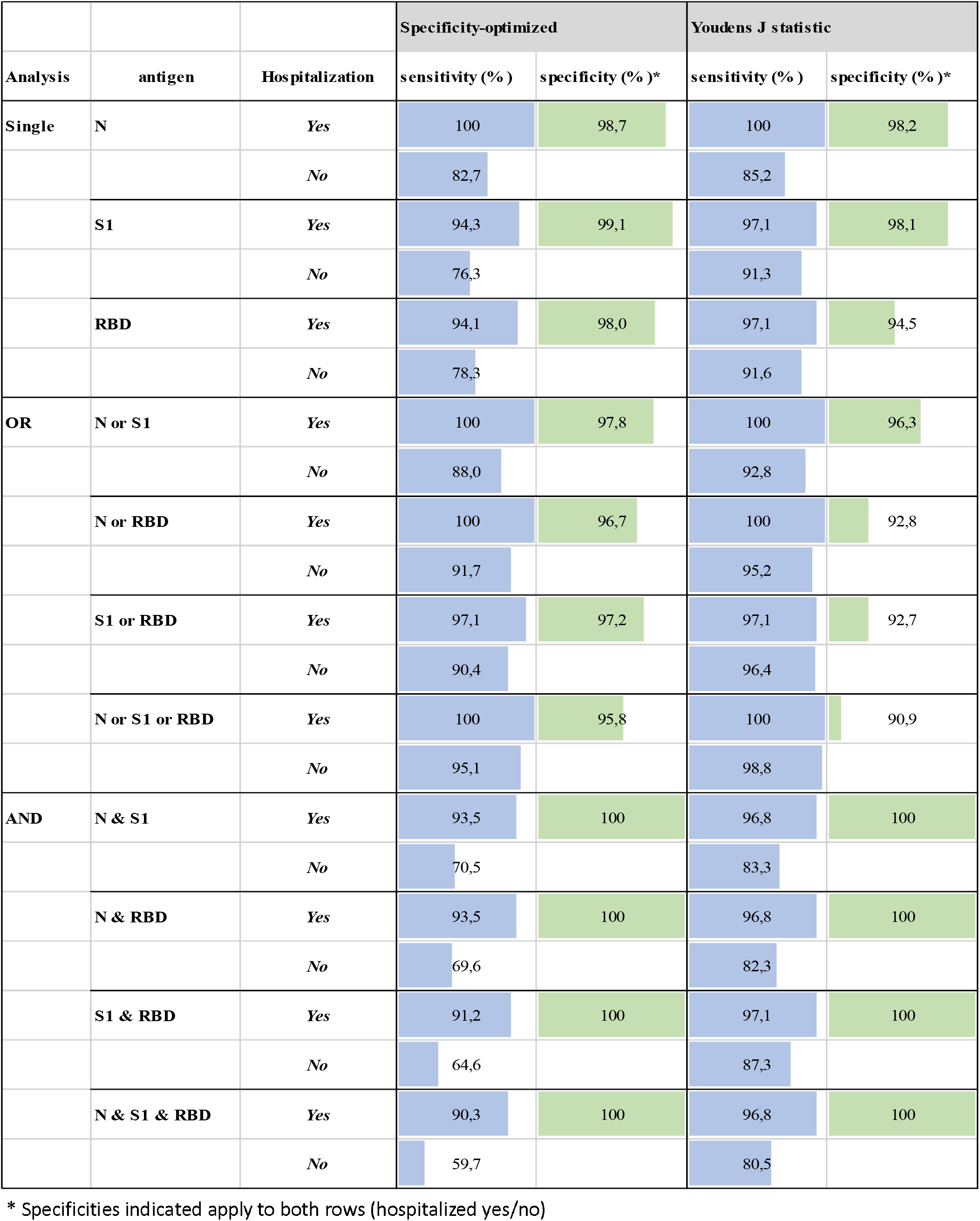
Specificity and sensitivity of single and multiplexed analyses of seroconversion.

### Sensitivity and specificity

Sera of 115 PCR-confirmed COVID-19 patients after 10 days of symptoms were tested in the assay and the results compared to a control panel of 408 control sera collected prior to the outbreak of SARS-CoV-2. In COVID-19 patients high concentrations of IgG were observed to all 3 antigens (**Figure 2A**). Despite clear discrimination of IgG concentrations between groups of control and COVID-19 patients, some samples overlapped between the two groups. Therefore, the specificity and sensitivity of the assay to discriminate between COVID-19 patients and controls using IgG concentrations was evaluated by an established statistical standard to analyze assay performance, the ROC analyses. For the ROC analyses hospitalized and non-hospitalized COVID-19 disease cases were included to provide a realistic evaluation of the performance of the assay. The area under the curves ranged from 0.9839-0.9859 (**Figure 2B**). The ROC generated cutoff concentrations of 14.8, 0.85 and 8.21 using the ROC Youden’s J statistic. To gain a higher specificity of the assay optimized for a low population seroprevalence, the cutoff concentrations were 19.7, 2.37 and 19.1 for N, S1 and RBD respectively (**Figure 2C**). The latter cutoffs resulted in a specificity of 98.5, 99.0 and 98.5 at a sensitivity of 89.4, 84.4 and 83.6 for N, S1 and RBD respectively.

**Figure 2.**
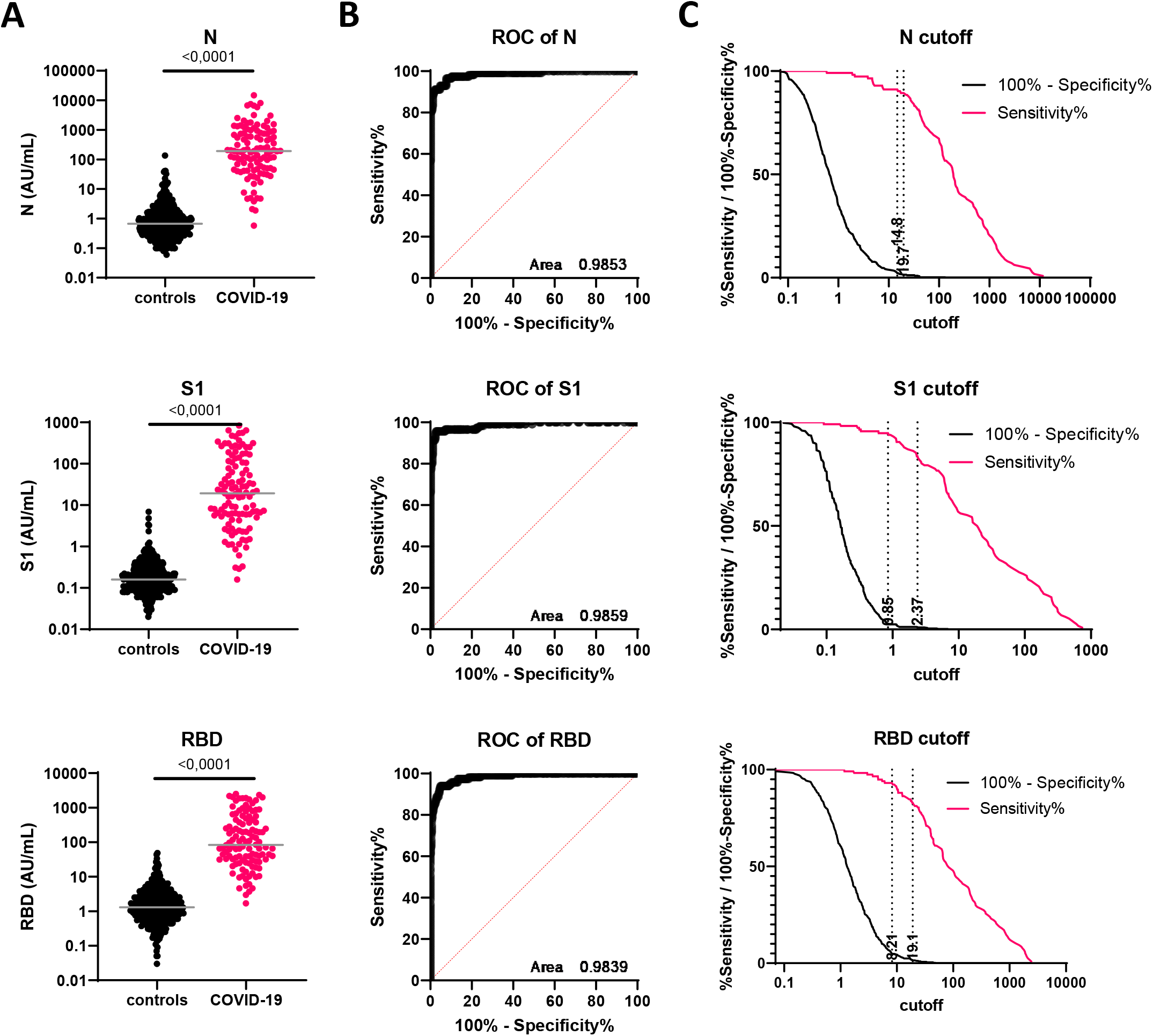
Ability of the assay to identify COVID-19 patients. **A**. 408 control sera and 115 COVID-19 sera collected after day 10 of symptoms were tested and compared for concentrations of IgG. **B**. the sera tested in A were analyzed by ROC. **C**. The ROC data were used to determine a Youden’s J statistic cutoff (lower cutoff) and a specificity-optimized cutoff of at least 98.5% specificity (higher cutoff).

### IgG in non-SARS-CoV-2 infections and SARS-CoV-2 infections of varying severity

To study how our assay discriminates between antibodies of individuals with varying etiology, antibodies were measured in a cross-sectional population panel (N=224), a panel of non-corona influenza-like illness (Non-hCoV ILI) patients (N=74) and non-SARS-CoV-2 corona hCoV ILI patients (N=110) and compared to PCR-confirmed COVID-19 patients’ samples. Part of the COVID-19 patients were enrolled into the hospital (N=50) because of a severe COVID-19 and those were compared to non-hospitalized COVID-19 cases (N=65). For each of the three negative control groups the majority of the samples reported concentrations below the cutoff for all three antigens (**Figure 3**). The number of false-positive samples ranged from 5-6 out of 404 or 408 samples tested for the different antigens. The Non-hCoV and hCoV ILI panels were from persons infected with multiple different non-SARS viruses including 4 different endemic coronavirus infections (**Table S2**). The proportion of false-positives did not increase by testing the convalescent sera from patients with a laboratory (PCR-) confirmed infection with either of the 4 seasonal coronaviruses (**Figure 3**, and data not shown), indicating that the antigens used in the assay are very selective for SARS-CoV-2 induced antibodies. Comparison of PCR-confirmed SARS-CoV-2 patients samples shows that all hospitalized patients induced antibodies to N and the majority of hospitalized patients induced antibodies to S1 and RBD. The majority of the non-hospitalized cases show antibody concentrations above the cutoff for N, whereas around 10% of the non-hospitalized patients did not produce antibodies above the cutoffs for S1 and RBD. Overall, the concentrations of antibodies in serum samples from patients that were hospitalized were significantly higher compared to patients that were not hospitalized.

**Figure 3.**
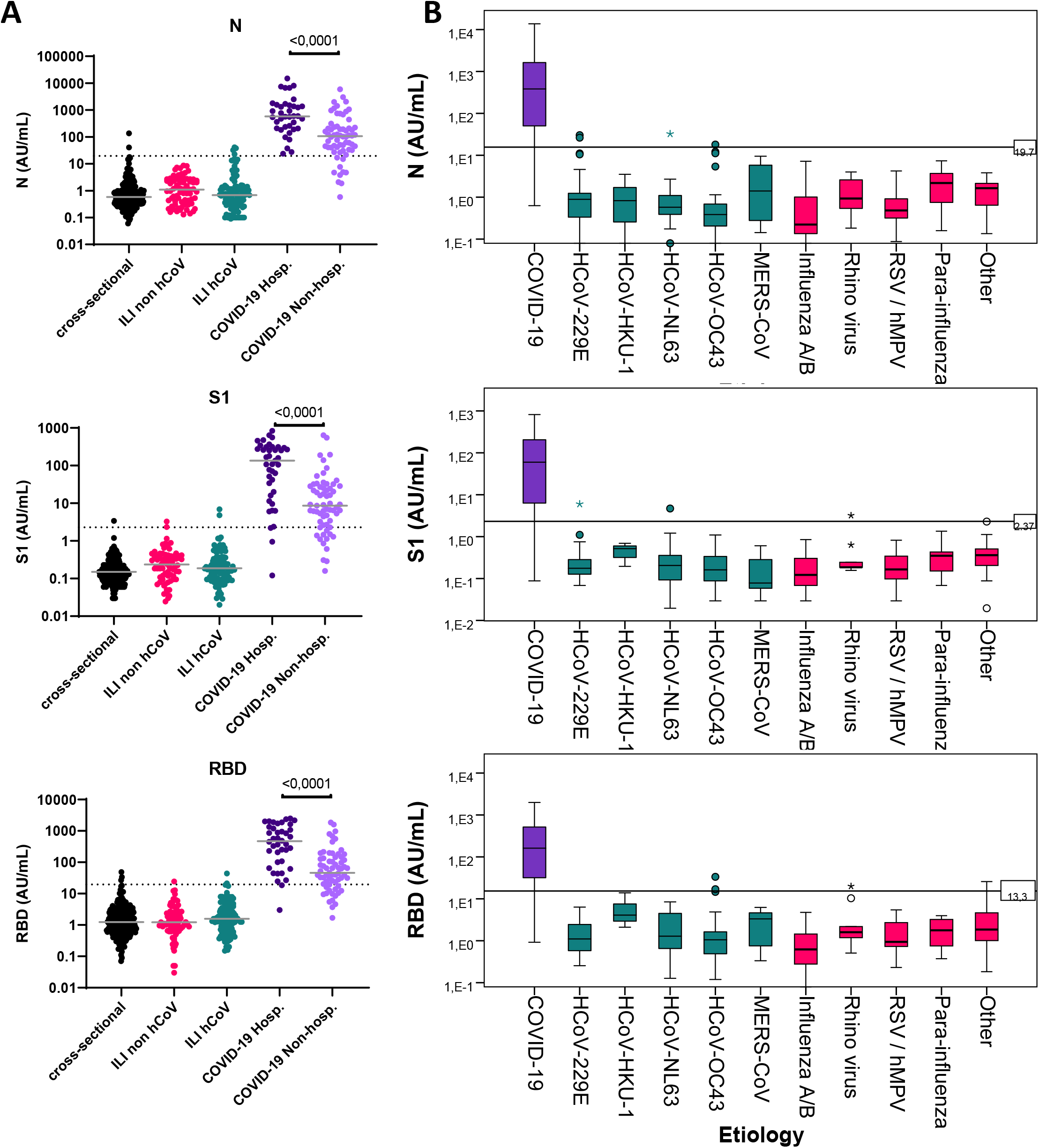
Discrimination of COVID-19 patients with varying severity from cross-sectional and ILI patients. **A**. 224 cross-sectional individuals ages 3-90 years, 75 ILI patients with non-coronavirus (non hCoV) and 109 non-SARS-CoV-2 ‘seasonal’ coronavirus infected ILI patients were compared to hospitalized and non-hospitalized COVID-19 patients. Statistical results (adjusted p-values of Tukey’s multiple comparison) between the COVID-19 groups are shown. B. Detailed etiology (see Table S2) and concentration data of ILI patients are shown to confirm that the assay discriminates SARS-CoV-2-specifici antibodies from antibodies induced by varying etiology.

### Kinetics of seroconversion

Following infection, an immune response is initiated, resulting in the production of serum antibodies. To study the time between onset of disease symptoms and the development of antibodies, paired serum samples were collected from the majority of patients. Data were separated for patients that were either admitted to the hospital or not (**Figure 4**). Apart from the paired samples from 2 patients that were obtained before 7 days after onset of disease, all other hospitalized cases show seroconversion for all three antigens tested (**Figure 4A**). In line with other reports, hospitalized COVID-19 patients seroconverted around day 10 of disease onset. Of 53 non-hospitalized cases, 48 seroconverted, whereas 5 showed slight increases in concentrations but failed to formally cross the cutoff value for any of the three analytes to be regarded a specific seroconversion. Hospitalized patients reached a plateau of antibody production shortly after two weeks of onset of symptoms, which took at least 25 days for the non-hospitalized cases (4-10 fold lower slope). As a consequence of the slower increase of antibody concentrations the time to detectable antibodies is delayed, especially with respect to antibodies reacting to S1 and RBD. The variance in the non-hospitalized cases is high compared to the hospitalized cases, which is illustrated by the lower R squares of the non-linear least square fit of the two patients groups.

**Figure 4.**
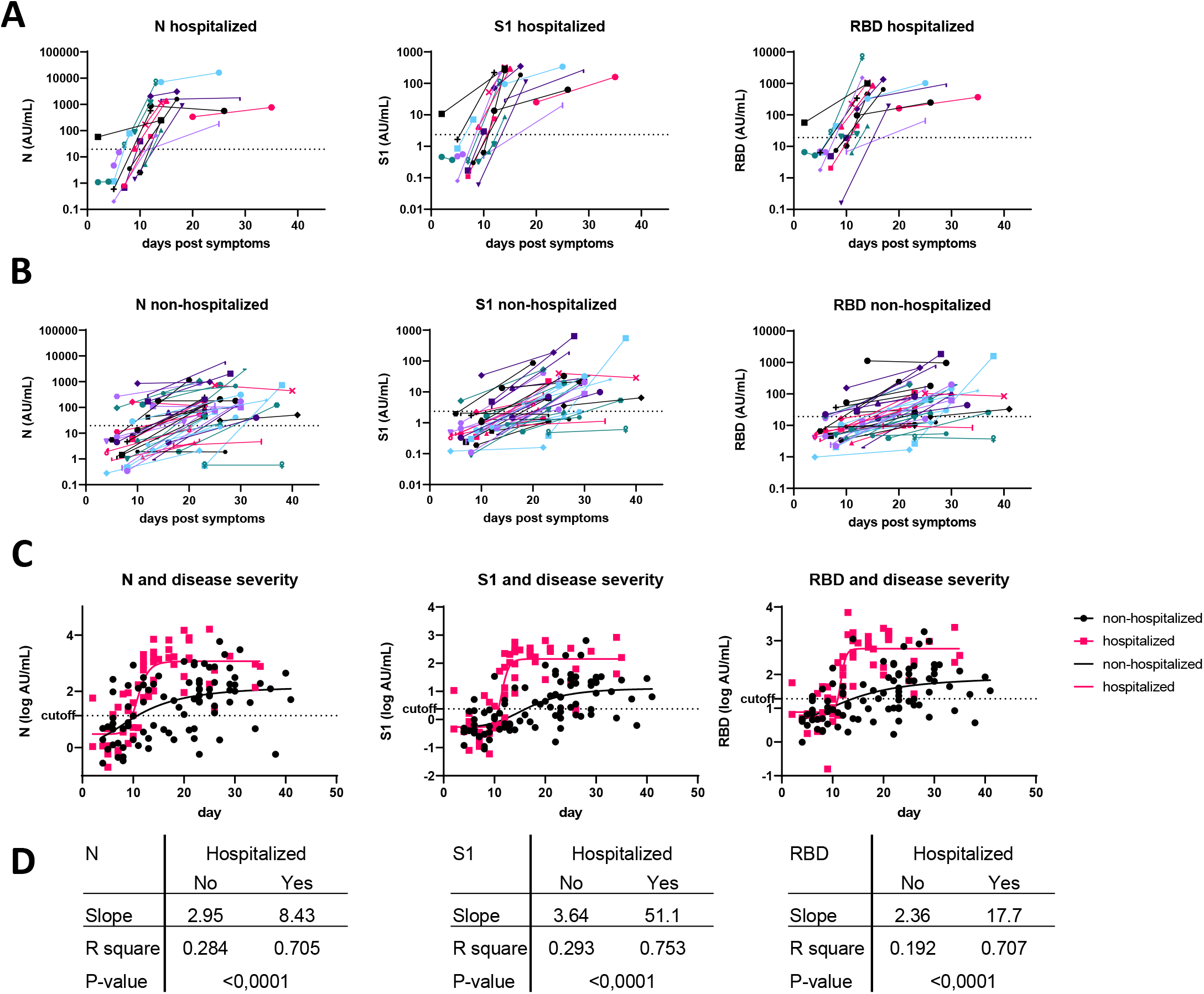
Kinetics of antibody production after disease onset in hospitalized and non-hospitalized COVID-19 patients. Paired samples were analyzed to identify changes in IgG concentrations in hospitalized **(A)** and non-hospitalized **(B)** COVID-19 patients. **C**. The log-transformed concentration data of the samples shown in A and B were fitted with a 4-parameter non-linear least squared fit. **D**. Of each patient with paired samples available one sample was selected randomly and data were fitted to estimate the slope, R square of the fits and the difference between the fitted lines determined.

### Multiplexed evaluation of seroconversion of different severity of COVID-19

The engagement of different structural SARS-CoV-2 proteins in one serological determination (multiplex testing) instead of one protein could improve the sensitivity and the specificity. If only one analyte is analyzed, the sensitivity for hospitalized cases is 94.1, 94.3 and 100% for RBD, S1 and N, respectively using the specificity-optimized cutoff (**Table 1**). Using the ROC Youden’s J statistic cutoff this sensitivity is 97.1% for both S1 and RBD and 100% for N. Non-hospitalized cases typically have lower concentrations of IgG, which reduces sensitivity; 76.3% for S1, up to 82.7 % for N using the specificity-optimized cutoff. Using the Youden’s J statistic cutoff, the sensitivity increases to 91.3% for S1.

In this multiplex approach an increased sensitivity can be obtained by evaluating a sample as positive when either one of the antibody concentrations determined is higher than the set cutoff (logical **‘OR’** analysis in Table 1). Any combination of antigen reaches a sensitivity of 100% when N is used in hospitalized cases and ranges from 90.4 (S1 or RBD) up to 95.1% (N or S1 or RBD) using the specificity-optimized cutoff. Applying the Youden’s J statistic cutoff results in a sensitivity for non-hospitalized cases of at least 92.8% (N or S1), up to 98.8% (N or S1 or RBD). The specificity of the Youden J analyses using N or S1 or RBD drops to 90.9%. This specificity is far too low for serosurveillance purposes in cases of low-prevalence. The specificity-optimized cutoff (95.8-97.8) is clearly better, which may be considered adequate if the true prevalence in the population is above 20%. Because in most countries the overall COVID-19 seroprevalence is currently under 10%, high specificity is required to provide reliable seroprevalence estimates (illustrated in **Table S1**). This could be achieved by defining a sample positive when at least two antibody test results in multiplex are above the cutoff. This resulted in a specificity of 100% for any of the combinations and both the specificity-optimized and the Youden’s J statistic-determined cutoffs (**Table 1**, logical **‘AND’**). As expected this increased specificity comes at the expense of the sensitivity. Here, if only S1 and RBD are taken into consideration, this combination resulted in the highest possible sensitivity of 87.3% and 97.1% for non-hospitalized and hospitalized patients respectively.

## DISCUSSION

We aimed to develop a high throughput quantitative assay to measure true concentrations of antibodies to Spike S1, Spike RBD and N of SARS-CoV-2. The assay presented here uses very little sample volume, which can be obtained from e.g. fingerstick blood, while retaining highly quantitative output. This bead-based multiplex immunoassay generates robust results and is able to discriminate COVID-19 with different degrees of disease severity, especially from day 10 of disease onward. The results of the assay presented here provide detailed insight into the performance of the assay in terms of parallelism between the references and sera containing different concentrations of antibodies. In addition, we show consistency of assay results when the same samples are measured on independent days, by different investigators using different batches of reagents; basically incorporating all potential variability.

Large population studies are in high demand to provide insight into the spread of the virus, the protective status of the population which can be used for policy makers to manage the pandemic or lift the lock-down measures [2, 3, 11, 12]. Assays have to generate accurate results to generate reliable seroprevalence data of the general population. In addition to knowing the performance of an assay we need to understand how the majority of infections in the general population relate to the induction of detectable antibodies. Our data comparing hospitalized and non-hospitalized COVID-19 antibody concentrations revealed that milder disease results in both lower levels of antibodies and later seroconversion, which is in line with previous reports [19, 20]. Moreover, some 10% of the non-hospitalized cases in our selection did not show any seroconversion at all, indicating that such mild infections may not be detected by serological assays.

Essential performance characteristics of assays aiming to identify seroprevalence in the population are the specificity and sensitivity. The specificity and sensitivity determine the positive and negative predictive value (PPV/NPV) of the assay given the prevalence of seropositivity in the population [21]. In current low prevalence settings insufficient specificity will generate a low PPV, resulting in a significant overestimation of the proportion of seropositive individuals (illustrated in **Table S1**). However, the accuracy of the reported sensitivity and specificity of an assay also highly depends on the patient selection used for this evaluation. E.g., using sera of severe COVID-19 patients will result in beneficial statistics of an assay, because of an acknowledged higher antibody concentration and seroconversion rate [22]. These statistics will not apply in a population serosurvey where the majority of persons will not develop severe COVID-19. For this reason, we included a heterogeneous group of COVID-19 patients’ samples, consequently reducing sensitivity. Scoring samples positive if at least 2 of the analytes generated positive results improved the specificity of the assay to 100% at a sensitivity > 90%. At a true seroprevalence of 5%, this would provide a seroprevalence estimate of 4.5%, and therefore will be much more accurate than using a single analyte. We recommend transparent reporting of underlying assay performance using heterogeneous panels of controls and COVID-19 patients. Furthermore, implementation of international reference materials as being distributed by e.g. NIBSC to facilitate comparison of seroepidemiological data between studies and countries is greatly recommended [1, 23].

From an immunological point of view, it needs to be established which SARS-CoV-2-specific antibodies correlate with protection. Antibodies to RBD of S1 have been shown to associate with neutralization of the virus in vitro, and preliminary data indicates that the antibodies reported in our assay correlate quantitatively with virus neutralization in vitro as well [6]. The data presented here show detection of total IgG. Other studies show that IgG subclasses are not equally induced by SARS-CoV-2 infection, with a bias towards the production of IgG3, at least in the first weeks after infection [24]. Infection with SARS-CoV-2 also induces the production of IgA and IgM, which can contribute to protection and in vitro neutralization of the virus, but is not detected in the data presented here and may enhance the sensitivity of the assay [7, 8, 25]. Follow-up studies need to establish the longevity of the production of antibodies, the degree of protection antibodies confer through various Fc receptor-mediated and other mechanisms and how B cell memory is induced. Such studies should also consider different viral loads detected in a patient and degree of severity of COVID-19.

In conclusion, we developed a robust multiplex assay to detect antibodies to SARS-CoV-2 in small blood volumes. Our study is unique in validating the assay against hCoV and Non-hCoV ILI panels. Because of the differences in seroconversion rates and quantitative antibody concentrations among non-hospitalized COVID-19 cases, which represents the majority of patients in the general population, further investigation is required to further improve assay for serosurveys. We show the advantages of multiplexed analysis in determining seroconversion and provide a framework for reliable seroprevalence estimates in different settings.

## Data Availability

Data referred to in the manuscript are available form corresponding authors upon reasonable request.

## ACKNOWLEDGEMENTS

The authors acknowledge Jorgen de Jonge and Puck van Kasteren for critically reviewing our manuscript and Gert-Jan Godeke for providing technical assistance.

